# Knowledge, Attitudes, and Practices of Advance Care Planning in Patients with Neurosurgical Conditions and Their Caregivers

**DOI:** 10.64898/2026.11.12.26367794

**Authors:** Vinh Anh Huynh, Mervyn Jun Rui Lim, Si En Ong, Ching Yi Lim, Gerald Choon Huat Koh, Pin Sym Foong

## Abstract

**Introduction:** Advance care planning (ACP) enables quality end-of-life (EOL) care that reflects patients’ values and preferences. However, ACP remains limited among individuals with neurosurgical conditions – such as stroke, glioma, or metastatic brain tumours – who often experience rapid decline and short prognoses. When ACP discussions occur only after cognitive deterioration or acute events, opportunities for meaningful conversations are lost. This study aims to assess the baseline knowledge, attitudes, and practices (KAP) of ACP among neurosurgical patients and their caregivers in Singapore.

**Methods:** We conducted a cross-sectional survey of 30 patients with symptomatic intracerebral haemorrhage (SICH) or brain tumours at the National University Hospital and their caregivers. Using validated ACP engagement questionnaires, we assessed participants’ KAP, and readiness on 5-point Likert scales (1 lowest - 5 best), alongside nine ACP knowledge statements.

**Results:** Patients demonstrated below-average ACP knowledge (mean = 4.1 / 9) and largely neutral attitudes (3.2 / 5) and perceptions (3.1 / 5). Misconceptions were common, including beliefs that ACP documents are legally binding, that preferred care cannot be delivered without ACP, and that ACP is solely a doctor’s responsibility. Consequently, both patients and caregivers showed low levels of ACP engagement.

**Conclusion:** The findings highlight the need to introduce ACP education and conversations earlier in the care journey, especially for individuals at risk of cerebrovascular or cardiovascular events. Early engagement, particularly among those with hypertension or other risk factors, presents a crucial window for preparing patients and families for future care decisions.

**CLINICAL IMPACT:** *What is New:*

- This study is the first to assess the knowledge, attitudes, and practices (KAP) of ACP among neurosurgical patients and their caregivers in Singapore.
- Findings revealed low levels of ACP engagement, below-average knowledge of and prevalent misconceptions regarding ACP among both patients and caregivers.

*Clinical Implications:*

- The findings highlight the need to introduce ACP education and conversations earlier in the care journey, especially for individuals at risk of cerebrovascular or cardiovascular events, such as those with hypertension or other risk factors.

## Background and introduction

### Delivering quality end-of-life care is increasingly becoming vital in the health systems and gaining more focus and priority

Advance care planning allows for better, quality EOL care that is aligned with patients values, priorities, preferences. Research has shown that engagement in ACP leads to better patient and caregiver outcomes, better care satisfaction, more goal-aligned care whereas The lack of ACP can result in confusion and anxiety for surrogates tasked with making critical decisions under time pressure.[1-3]

Patients with neurosurgical conditions face poor and often short prognoses, yet little is known about their knowledge of, or needs for, advance care planning (ACP). Median survival is approximately 18 months for glioma[4], 6 – 9 months for metastatic brain tumours,[5] and 6 – 12 months for haemorrhagic stroke patients,[6] making it crucial to introduce ACP at diagnosis. Hence, timely ACP is especially challenging in this group. When discussions occur only after sudden deterioration such as with brain trauma or stroke, opportunities for meaningful values elicitation and informed conversations may be lost.[7, 8] For this reason, early initiation of ACP—ideally before mental capacity is compromised or before acute episodes occur—is more urgent for patients with neurosurgical conditions than for those with illnesses that typically allow for longer prognoses. In addition, family participation and engagement are critical. Evidence suggests that dyadic approaches, in which patients and families are involved together, are associated with better end-of-life preparedness and more positive experiences.[9, 10] However, caregivers in Singapore, and in Asian cultures in general, prioritize family-centred decision-making and are less likely to be open to EOL discussions making meaningful dialogue about end-of-life preferences much harder as compared to those in western countries where the emphasis on individual autonomy and direct conversations about advance care planning make it easier for caregivers and patients to explore EOL options.[11]

As such, before promoting improved knowledge, attitudes, and engagement (KAP) of ACP among this population of patients and caregivers, we conducted a cross-sectional study to better understand the baseline ACP KAP among this population of elevated risk of sudden death or incapacitation. These findings will help complement previous work in exploring ACP KAP among healthcare workers (HCW) in neurology, neuro□oncology and surgery in Singapore and have potential to inform policies to enhance ACP education and promote uptake of ACP conversations among this particularly vulnerable population.[12]

## Methods

### Study setting and recruitment

We recruited adult patients with glioma, brain metastases or Symptomatic Intracerebral Haemorrhage (SICH) managed by ***[redacted for peer review]***. The inclusion criteria were: (a) patients with a histological diagnosis of glioma or metastatic brain disease; (b) patients aged 21 and above; and (c) patients able to communicate in English or Chinese. Patients who are unable to participate due to poor cognition would be excluded from this study. We also recruited caregivers caring for eligible patients using the following inclusion criteria were: (a) caring for patients with the diagnosis of patients inclusion criteria; (b) aged 21 and above; and (c) able to communicate in English or Chinese. Written informed consent was obtained for all participants. This study was conducted in accordance with the principles of the Declaration of Helsinki and approved by ***[redacted for peer review]***.

### Data collection

A standardized questionnaire on KAP of ACP was developed using (a) validated questionnaires from the international literature; (b) concepts of Theory of Planned Behaviour; and (c) in consultation with a multi-disciplinary panel of specialists involved in the care of patients with neurological diseases.[12] For patients, the questionnaire measured the following domains: ACP knowledge (9 items), attitudes (16 items), and perception (16 items), adapted from Zhou et al.[13] Knowledge of ACP items contained nine statements allowing respondents to choose one appropriate answer (True, False, or Don’t Know) for each statement (**Supplementary Table 1**). ACP attitude and perception items used a five-point Likert scale ranging from 1 (strongly agree) to 5 (strongly disagree). ACP attitudes were measured across four subthemes: 1) ACP improves EOL care (2 items), 2) ACP is detrimental to patients (5 items), 3) ACP is a personal responsibility (4 items), and 4) ACP importance in EOL decision-making (5 items). Higher scores indicate more positive attitude and perception of ACP. Patient participants’ ACP engagement was measured using the 4-item ACP engagement instrument.[14] The instrument measures respondents’ readiness to engage in ACP behaviours including: 1) signing official paper naming nominated healthcare spokesperson, 2) talking to medical decision maker and 3) medical providers about care goals and wishes, and 4) formally documenting care goals and wishes. Responses for each ACP engagement behaviours also used a 5-point Likert scale, ranging from “I have never thought about it” to “I have already done it” with higher scores indicate greater ACP engagement.

**Table 1.** Demographic and health descriptive statistics.

| **Mean (SD) / n (%)** | **Patients**<br>**N = 15** | **Caregivers**<br>**N = 15** |
|---|---|---|
| *Sociodemographic* |  |  |
| **Gender, n(%)** |  |  |
| Male | 11 (73.3%) | 5 (33.3%) |
| Female | 4 (26.7%) | 9 (60.0%) |
| **Age** | 59.0 (12.4) | 52.0 (19.9) |
| **Race** |  |  |
| Chinese | 11 (73.3%) | 9 (60.0%) |
| Indian | 0 (0%) | 4 (26.7%) |
| Malay | 4 (26.7%) | 1 (6.7%) |
| Others | 0 (0%) | 0 (0%) |
| **Religion** |  |  |
| Buddhism | 8 (53.3%) | 4 (26.7%) |
| Christianity | 1 (6.7%) | 3 (20.0%) |
| Hinduism | 0 (0%) | 0 (0%) |
| Islam | 4 (26.7%) | 5 (33.3%) |
| No religion | 0 (0%) | 2 (13.3%) |
| Others | 2 (13.3%) | 0 (0%) |
| **Education** |  |  |
| Secondary and below |  | 7 (46.7%) |
| A-Levels or Diploma |  | 4 (26.7%) |
| Bachelor’s Degree |  | 3 (20.0%) |
| Post-graduate (Master’s or PhD) |  | 0 (0%) |
| **Household income per month** | 2430 (2600) | 3990 (2750) |
| **Housing** |  |  |
| HDB/JTC Flat (1-2 room) | 0 (0%) | 0 (0%) |
| HDB/JTC Flat (3-4 room) | 8 (53.3%) | 4 (26.7%) |
| HDB/JTC Flat (5 room and above/ Exec) | 4 (26.7%) | 7 (46.7%) |
| Condominium/ private apartment | 3 (20.0%) | 2 (13.3%) |
| Landed | 0 (0%) | 0 (0%) |
| **Marital status** |  |  |
| Single | 1 (6.7%) | 5 (33.3%) |
| Married | 13 (86.7%) | 9 (60.0%) |
| Divorced / Widowed | 1 (6.7%) | 0 (0%) |
| **Relationship to patient (‘Patient is my…')** |  |  |
| Mother/Father |  | 4 (26.7%) |
| Grandmother/Grandfather |  | 0 (0%) |
| Wife/Husband |  | 9 (60.0%) |
| Sibling |  | 1 (6.7%) |
| *Health-related measures* |  |  |
| **Diagnosis of patient** |  |  |
| Glioma | 2 (13.3%) | 1 (6.7%) |
| SICH / stroke | 13 (86.7%) | 14 (93.3%) |
| **Recruitment site** |  |  |
| Inpatient | 4 (26.7%) |  |
| Outpatient | 11 (73.3%) |  |
| **Self-rated health rating (range: 0 - 100)** | 49.4 (21.0) | 81.1 (14.6) |
| **Health-related quality of life (range: 0 - 1)** | 0.425 (0.525) | 0.889 (0.168) |
| **Comorbidities** |  |  |
| Hypertension | 12 (80.0%) |  |
| High cholesterol | 9 (60.0%) |  |
| Diabetes mellitus | 5 (33.3%) |  |
| **Caregivers Quality of Life Scale (range: 0 – 100)** |  | 76.3 (16.0) |
| Physical Wellbeing |  | 92.5 (17.6) |
| Mental Wellbeing |  | 70.0 (31.3) |
| Experience and Meaning |  | 65.0 (32.1) |
| Impact on Daily Life |  | 80.0 (19.9) |
| Financial Wellbeing |  | 65.0 (28.0) |

**Notes:** SD – Standard deviation; SICH – Symptomatic Intracerebral Hemorrhage

For caregivers, we measured ACP engagement using the 17-item ACP surrogate decision making (SDM) scale.[15] The ACP-17 SDM assesses surrogate decision-makers’ (SDM) role in ACP via three factor domains (Serving as surrogate decision-maker – 7 items, Contemplation – 4 items, and Readiness – 6 items) where each item is scored on a 5-point Likert scale. Higher ACP-17 SDM scores indicate greater level of SDM’s preparedness to make future medical decisions on behalf of a loved one.

We also collected demographic details such as age, gender, race, religion, health-related quality of life and prior medical diagnoses. Clinical data related to the patient’s tumour and the management of their tumour was obtained from electronic medical records. The English version of the patient survey questionnaire was also translated into Chinese using validated questionnaires from the international literature or by the research team in accordance to best practice.[16] The full questionnaires are shown in **Supplemental Appendix Survey Instrument**.

## Results

### Descriptive statistics

Between May 2024 and June 2025, 15 patients and 15 caregivers consented to and completed the surveys, yielding a response rate of 14%. Characteristics of patient and caregiver respondents are presented in **Table 1**. Among patient participants, the mean age was 59.0 ± 12.4, most were male (73.3%), Chinese (73.3%), married (86.7%) , diagnosed with SICH (86.7%), recruited from outpatient setting (73.3%), and have underlying comorbidities such as hypertension (80.0%) and high cholesterol (60.0%). Among caregiver participants, the mean age was 52.0 (SD: 19.9) and most were female (60.0%), married (60.0%), Chinese (60.0%), and caring for their spouse (60.0%), followed by parents (26.7%).

### ACP experience, knowledge, attitude and practice

#### Among patients, none reported having made an ACP or knowing a family or friend who have made an ACP

ACP knowledge score was below numerical average (mean: 4.07 ± 1.98 out of 9), with evidence indicating significant misconception or misunderstanding of ACP regarding several aspects of ACP **(Table 2**). For example, only one patient participant (6.7%) was able to accurately identified that ACP documents are not legally binding, and only four (26.7%) were able to identify that a person does not need an ACP before end-of-life wishes can be followed. On the other hand, most patient participants (87.8%) were aware that they could change their ACP at any time (**Supplementary Figure 1**).

**Table 2.**
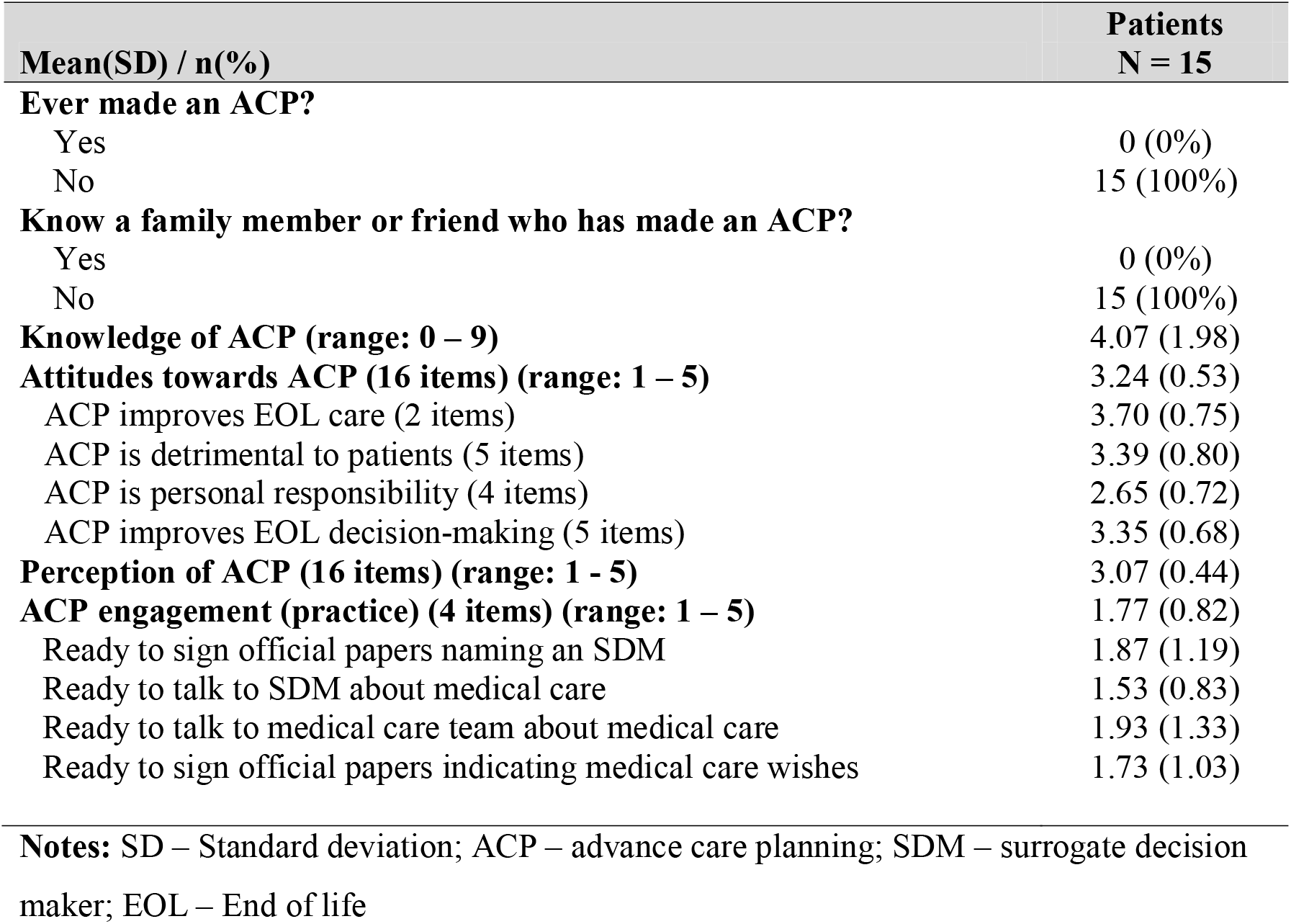
ACP experience, knowledge, attitude, perceptions, and readiness among patients.

Patients’ attitudes and perception of ACP were largely neutral (mean scores of 3.24 ± 0.53 and 3.07 ± 0.44 out of 5 respectively). Notably, most patient respondents scored below average for the personal responsibility subscale of ACP attitudes, suggesting strong hesitancy to take the initiative to bring up ACP unless suggested by their care team with close to half of the respondents “strongly agree” that “ACP is doctor or healthcare team’s responsibility.”

Last, we found very poor ACP engagement/readiness among patients, where most have yet to think about care planning at all (mean score 1.77 ± 0.82 out of 5) (**Table 2**). Of note, patient respondents reported the lowest score for the item “…ready to talk to SDM about the kind of medical care” they would want (mean: 1.53 ± 0.83), highlighting significant barriers or taboo on the patient’s side, surrounding discussing EOL care and values with SDMs.

**Among caregivers, only two caregivers (13.3%) have been involved in making end-of-life decisions for someone else, although it was not possible to verify if such involvement was in the formal ACP context, and only three out of 15 reported that the patient has ever expressed their wishes or preferences about their care at the end-of-life (20.0%)** The mean score for the ACP-17 SDM domains were 3.59 ± 1.17, 2.63 ± 1.57, and 2.59 ± 1.46 out of 5 for the serving as surrogate decision-maker, contemplation, and readiness domains respectively. The overall ACP-17 SDM score is average, at 3.03 ± 1.16 out of 5 (**Table 3**), which is similar to the results found from a larger population of patients with serious illness.**[17]**

**Table 3.** ACP experience, knowledge, attitude, perceptions,and readiness among caregivers.

| **Mean(SD) / n(%)** | **Caregivers**<br>**N = 15** |
|---|---|
| **Been involved in making end-of-life decisions for someone else in the past?** |  |
| Yes | 2 (13.3%) |
| No | 12 (80.0%) |
| **Has a doctor given the patient a medical prognosis of 12 months or less left to live?** |  |
| Yes | 0 (0%) |
| No | 9 (60.0%) |
| Not sure | 5 (33.3%) |
| **Has the patient ever expressed their wishes or preferences about their care at the end-of-life?** |  |
| Yes | 3 (20.0%) |
| No | 7 (46.7%) |
| Not sure | 4 (26.7%) |
| **ACP engagement SDM (17 items) (range: 1 – 5)** | 3.03 (1.16) |
| Serving domain (7 items) | 3.59 (1.17) |
| Contemplation domain (4 items) | 2.63 (1.57) |
| Readiness domain (6 items) | 2.59 (1.46) |

**Notes:** SD – Standard deviation; ACP – advance care planning; SDM – surrogate decision maker; EOL – End of life

## Discussion

### The study aims to assess the level of knowledge attitude, perception and engagement of ACP among patients with neurosurgical disorders and their caregivers

We found that although patient participants demonstrated below-average knowledge of ACP, their attitudes and perceptions appeared largely neutral. One explanation could be that patients lacked sufficient understanding of ACP to form strong opinions. However, it is more likely that this neutrality reflects the impact of an ongoing, national outreach and education efforts at the time of data collection. These campaigns may have helped frame ACP in balanced—even positive—ways, preventing overly negative perceptions and reducing resistance to ACP among the general public.

There are significant misconceptions surrounding ACP, such as nonbinding nature of ACP or that preferred care cannot be delivered unless ACP is completed. There is also a (mis)perception that ACP is not a personal responsibility, which is consistent with past studies show low personal relevance and dependency on others, such as doctors or family, for planning and medical decision-making. Some individuals prefer professionals to make choices, expressing reluctance to take personal ownership of ACP, especially older adults.[18] This misconception can result in missed opportunities for autonomy and meaningful preparation for unforeseen health events.[18, 19] Given these factors, it is unsurprising that ACP engagement is significantly low, both among patients and caregivers, consistent with findings by Ng. et al., who found that only 14.4% of the Singapore general public had heard about ACP and had perception barriers surrounding starting ACP conversation.[20]

Given the gaps observed in knowledge and attitudes, this study argues that it is beneficial to introduce ACP education and conversations earlier in the patient journey for people at risk of cerebrovascular diseases, i.e. SICH and stroke, or cardiovascular, i.e. heart attack. A diagnosis of hypertension or fulfilling the elevated risk criteria is a major predictor of cerebro-and cardiovascular risk,[21] and indicates a potential window for earlier ACP engagement. Hypertension was present in 80% of our sample. While hypertension management should remain the clinical priority, incorporating gradual and early ACP discussions into care plans could help prepare patients for the elevated risk of sudden cardiovascular events. Achieving this, requires a careful design of customized messaging for patients with hypertension. Programs have to balance the challenge the negative connotations and misconceptions often linked with ACP. In many contexts, ACP is misinterpreted as a sign of hopelessness or equated with fatalism and taboos around death.[22, 23] Hence, promoting ACP risks *demotivating* the necessary preventive behaviours such as increasing exercise, improving diet and adhering to medications. Balancing these countering forces is critical to fostering greater acceptance and meaningful engagement with ACP among populations with hypertension who are at greater risk of sudden functional decline.

Our findings should be interpreted with several caveats. First, we faced significant challenges in recruiting newly diagnosed neurosurgical patients, who often present after an acute event and may already have cognitive impairments. As a result, our sample size is small and not generalisable to the wider population. It is also possible—though unlikely— that patients who declined participation or were excluded due to cognitive impairment may have had greater knowledge, attitudes, and practices regarding ACP. Future research should adopt a longitudinal design, particularly focusing on other subpopulations at elevated risk of neurosurgical conditions, such as individuals with hypertension but no previous cerebrovascular events. This would allow for a clearer understanding of how ACP knowledge and engagement evolve over time, and help identify optimal moments for introducing ACP education and support.

### Financial disclosure

This work was funded by ***[redacted for peer review]***. The funder has no part in study design; collection, management, analysis, and interpretation of data; writing of the report; and the decision to submit the report for publication.

## Supporting information

Supplementary Appendix Survey Instrument - Patients

Supplementary Appendix Survey Instrument - Caregivers

## Data Availability

All data produced in the present study are available upon reasonable request to the corresponding author

## Acknowledgement

The authors used ChatGPT-4o to assist with rewording and take full responsibility for the final reviewed and edited content.

