## Supplementary Materials for "Knowledge, Attitudes, and Practices of Advance Care Planning in Patients with Neurosurgical Conditions and Their Caregivers"

### **Supplementary material**

#### **Supplementary Table 1. ACP Knowledge questions and correct answers**

| **Statements** | **True** | **False** | **Don’t Know** |
| --- | --- | --- | --- |
| 1. ACP allows me to share my beliefs and values | √ |  |  |
| 2. ACP can be in written or oral form | √ |  |  |
| 3. ACP is an ongoing process and I can change my decisions anytime | √ |  |  |
| 4. ACP is a legally binding document |  | √ |  |
| 5. Only patients with 1 year left to live should be referred for ACP |  | √ |  |
| 6. A person needs an ACP before end-of-life wishes can be followed |  | √ |  |
| 7. A person with Legal Power of Attorney (LPA) has the authority to make health care decisions for a person when they cannot | √ |  |  |
| 8. A person can only appoint a family member for LPA |  | √ |  |
| 9. The healthcare team is obliged to provide aggressive treatment that a person or their family requests regardless of its likely success |  | √ |  |

#### **Supplementary Figure 1. ACP Knowledge questions and correct answers among patients (N = 15)**

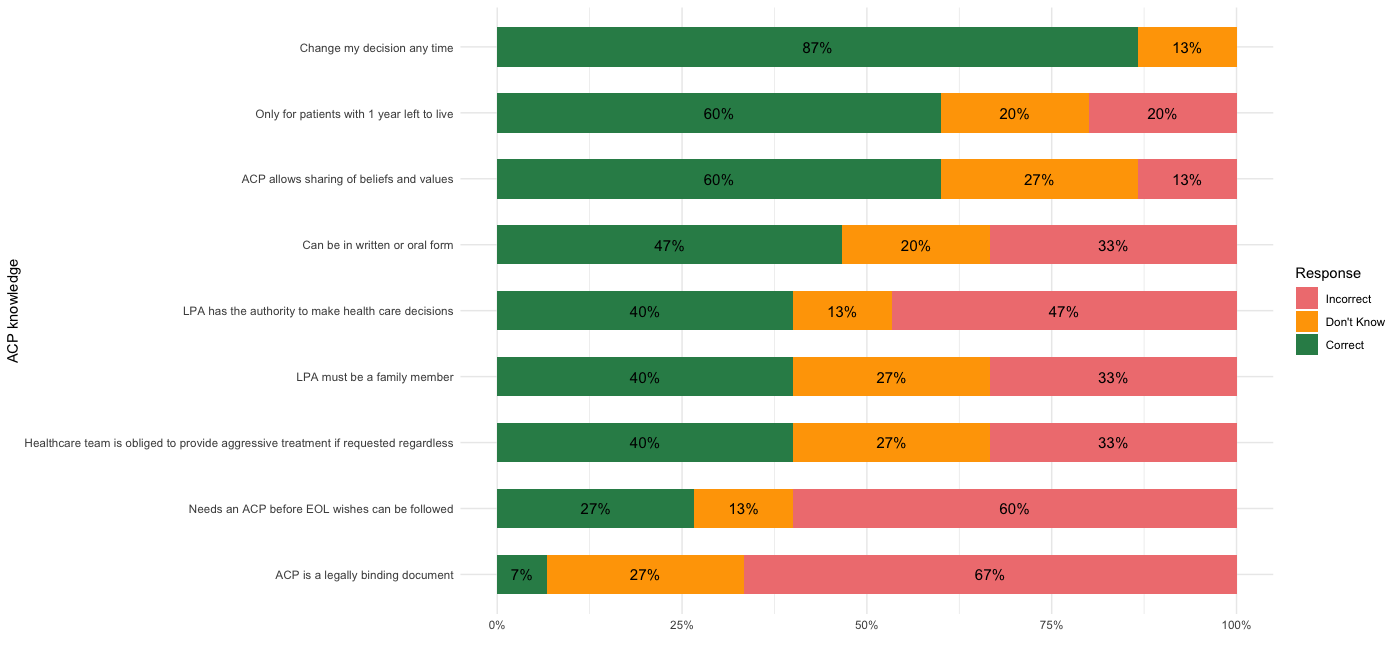

**Notes:** ACP – advance care planning; EOL – End of life; LPA – Lasting power of Attorney
