## Supplementary Appendix Survey Instrument - Patients for "Knowledge, Attitudes, and Practices of Advance Care Planning in Patients with Neurosurgical Conditions and Their Caregivers"

### **Supplementary Appendix Survey Instrument – Caregiver**

**ACP-NES Study Questionnaire for Patients**

Introduction

This study is titled “Advance Care Planning in Neurology, Neuro-oncology, and Surgery (ACP-NES Study)”. Thank you for taking the time to fill in this online survey. Before doing so, kindly read the consent form to ensure that you understand the study and what is involved.

[Consent form will be in hardcopy form.]

Section A: Socio-demographics

1. Gender

☐M

☐F

2. Race

☐Chinese

☐Indian

☐Malay

☐Others ___

3. Religion

☐Buddhism

☐Christianity

☐Hinduism

☐Islam

☐Others ___

4. Socio-economic status
   4.1. Income brackets
   4.2. Housing type
5. Marital status:

☐Single

☐Married

☐Divorced

☐Widowed

Section B: Experience [Suggested questions]

Have you ever made an ACP?

☐Yes

☐No

Do you know a family member or friend who has made an ACP?

☐Yes

☐No

Section C: Knowledge of ACP

ACP allows me to share my beliefs and values: **True**/False/Don’t know

ACP can be in written or oral form: **True**/False/Don’t know

ACP is an ongoing process and I can change my decisions anytime: **True**/False/Don’t know

ACP is a legally binding document: True/**False**/Don’t know

Only patients with 1 year left to live should be referred for ACP: True/**False**/Don’t know

A person needs an ACP before end-of-life wishes can be followed: True/**False**/Don’t know

A person with Legal Power of Attorney (LPA) has the authority to make health care decisions for a person when they cannot: **True**/False/Don’t know

A person can only appoint a family member for LPA: True/**False**/Don’t know

The healthcare team is obliged to provide aggressive treatment that a person or their family requests regardless of its likely success: True/**False**/Don’t know

Section D: Attitudes towards ACP

|  | Strongly Agree | Agree | Neutral | Disagree | Strongly disagree |
| --- | --- | --- | --- | --- | --- |
| **Theme: ACP improves EOL care** |  | | | | |
| ACP can improve patients’ and families’ satisfaction about end-of-life care. |  |  |  |  |  |
| ACP reduces the likelihood of futile treatment at the end of life. |  |  |  |  |  |
| **Theme: ACP is detrimental to patients** |  | | | | |
| Completing an ACP is emotionally draining. |  | | | | |
| ACP will speed up the dying process in many patients. |  |  |  |  |  |
| ACP destroys patients or their family members’ sense of hope. |  |  |  |  |  |
| Having an ACP will make my family feel left out of caring for me. |  |  |  |  |  |
| ACP means that the healthcare team is no longer pursuing active treatment for a patient. |  |  |  |  |  |
| **Theme: personal responsibility** |  | | | | |
| It is better to make an ACP when I am healthy. |  | | | | |
| I don’t know enough about healthcare to prepare an ACP. |  | | | | |
| ACP is my doctor or healthcare team's responsibility. |  |  |  |  |  |
| When there is a good doctor-patient relationship, ACP is not needed. |  |  |  |  |  |
| **Theme: EOL decision-making** |  | | | | |
| ACP is not necessary because use of life-sustaining treatments is a medical decision based on patients’ best interests. |  |  |  |  |  |
| ACP is not necessary because my family will know what to do when the time comes. |  |  |  |  |  |
| ACP is helpful to clarify patients’ goals and preferences for end-of-life care. |  |  |  |  |  |
| ACP can help to alleviate burden on family decision makers. |  |  |  |  |  |
| ACP can help to prevent disputes between health care team and family members on medical decisions. |  |  |  |  |  |

Section E: Perceptions of ACP

|  | Strongly Agree | Agree | Neutral | Disagree | Strongly disagree |
| --- | --- | --- | --- | --- | --- |
| I think that most patients with life-threatening illnesses, if asked, want to discuss their wishes for end-of-life care. |  |  |  |  |  |
| I feel comfortable discussing issues related to death and dying with my healthcare team. |  |  |  |  |  |
| Patients should be routinely screened for ACP referral. |  |  |  |  |  |
| It is not appropriate for my healthcare team to initiate ACP unless asked by myself or my family members. |  |  |  |  |  |
| I usually find end-of-life care discussion a taboo. |  |  |  |  |  |
| My family members may find it difficult to have end-of-life care discussion with myself and the ACP team. |  |  |  |  |  |
| I feel comfortable discussing issues related to death and dying with my family members. |  |  |  |  |  |
| If I have a documented ACP in my electronic health records, I feel that my doctor will strictly follow my stated wishes. |  |  |  |  |  |
| The existing ACP policy and guidelines is clear. |  |  |  |  |  |
| ACP should not be started before the patients’ condition worsens because their preferences may change according to the context. |  |  |  |  |  |
| It is difficult for me and/or my family members to reach consensus on end-of-life care. |  |  |  |  |  |
| If my wishes conflict with my family’s wishes, the doctor should comply with my wishes. |  |  |  |  |  |
| If the healthcare team thinks treatment is beneficial but I have indicated that I do not want treatment in my ACP, they should ignore or overrule my ACP. |  |  |  |  |  |
| Patients with decision-making capacity who are not terminally ill should have a right to refuse treatment even if that decision may lead to death. |  |  |  |  |  |
| I am ready to make an ACP even if it is difficult for my loved ones. |  |  |  |  |  |
| I think I may make an ACP in the next 6 months. |  |  |  |  |  |

**EQ5D**

**MOBILITY**

- ⊗I have no problems in walking about (1)
- ⊗I have slight problems in walking about (132)
- ⊗I have moderate problems in walking about (133)
- ⊗I have severe problems in walking about (134)
- ⊗I am unable to walk about (135)

**SELF-CARE**

- ⊗I have no problems washing or dressing myself (1)
- ⊗I have slight problems washing or dressing myself (6)
- ⊗I have moderate problems washing or dressing myself (7)
- ⊗I have severe problems washing or dressing myself	(8)
- ⊗I am unable to wash or dress myself (9)

**USUAL ACTIVITIES (e.g. work, study, housework, family or leisure activity):**

- ⊗I have no problems doing my usual activities (6)
- ⊗I have slight problems doing my usual activities (7)
- ⊗I have moderate problems doing my usual activities (8)
- ⊗I have severe problems doing my usual activities (9)
- ⊗I am unable to do my usual activities (10)

**PAIN / DISCOMFORT**

- ⊗I have no pain or discomfort (1)
- ⊗I have slight pain or discomfort (6)
- ⊗I have moderate pain or discomfort (7)
- ⊗I have severe pain or discomfort (8)
- ⊗I have extreme pain or discomfort (9)

**ANXIETY / DEPRESSION**

- ⊗I am not anxious or depressed? (1)
- ⊗I am slightly anxious or depressed? (6)
- ⊗I am moderately anxious or depressed? (7)
- ⊗I am severely anxious or depressed? (8)
- ⊗I am extremely anxious or depressed? (9)

| Page Break |
| --- |

We would like to know how good or bad your health is TODAY.

This scale is numbered from 0 to 100

**100** means the best health you can imagine.

**0** means the worst health you can imagine.

Mark an X on the scale to indicate how your health is TODAY.

|  | **The worst health you can imagine** | **The best health you can imagine** |
| --- | --- | --- |

|  | 0 | 5 | 10 | 15 | 20 | 25 | 30 | 35 | 40 | 45 | 50 | 55 | 60 | 65 | 70 | 75 | 80 | 85 | 90 | 95 | 100 |
| --- | --- | --- | --- | --- | --- | --- | --- | --- | --- | --- | --- | --- | --- | --- | --- | --- | --- | --- | --- | --- | --- |

| () | 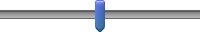 |
| --- | --- |

**Depression, Anxiety and Stress Scale - 21 Items (DASS-21)**

|  | Did not apply to me at all | Applied to me to some degree, or some of the time | Applied to me to a considerable degree or a good part of time | Applied to me very much or most of the time |
| --- | --- | --- | --- | --- |
| 1. I found it hard to wind down | 1 | 2 | 3 | 4 |
| 2. I was aware of dryness of my mouth | 1 | 2 | 3 | 4 |
| 3. I couldn’t seem to experience any positive feeling at all | 1 | 2 | 3 | 4 |
| 4. I experienced breathing difficulty (e.g. excessively rapid breathing, breathlessness in the absence of physical exertion) |  |  |  |  |
| 5. I found it difficult to work up the initiative to do things |  |  |  |  |
| 6. I tended to over-react to situations |  |  |  |  |
| 7. I experienced trembling (e.g. in the hands) |  |  |  |  |
| 8. I felt that I was using a lot of nervous energy |  |  |  |  |
| 9. I was worried about situations in which I might panic and make a fool of myself |  |  |  |  |
| 10. I felt that I had nothing to look forward to |  |  |  |  |
| 11. I found myself getting agitated |  |  |  |  |
| 12. I found it difficult to relax |  |  |  |  |
| 13. I felt down-hearted and blue |  |  |  |  |
| 14. I was intolerant of anything that kept me from getting on with what I was doing |  |  |  |  |
| 15. I felt I was close to panic |  |  |  |  |
| 16. I was unable to become enthusiastic about anything |  |  |  |  |
| 17. I felt I wasn’t worth much as a person |  |  |  |  |
| 18. I felt that I was rather touchy |  |  |  |  |
| 19. I was aware of the action of my heart in the absence of physical exertion (e.g. sense of heart rate increase, heart missing a beat) |  |  |  |  |
| 20. I felt scared without any good reason |  |  |  |  |
| 21. I felt that life was meaningless |  |  |  |  |

**PREPARATION ENGAGEMENT SURVEY**

**We will ask about your experiences and opinions.** We may ask about things that you have already done, or have not thought about at all. Just answer as honestly as you can.

***Over the next few sections we will be asking you about 2 topics:***

1. Medical decision makers, or surrogates

2. Deciding what matters most in life

ACP4_1 **1. Medical Decision Makers** The following question asks about medical decision makers. A medical decision maker is a family member or friend who can make decisions for you if you were to become too sick to make your own decisions.

**There are no right or wrong answers. We greatly appreciate your honest opinions.**

**1. How ready are you to SIGN OFFICIAL PAPERS naming a person or group of people to make medical decisions for you?**

- I have never thought about it (1)
- I have thought about it, but I am not ready to do it (2)
- I am thinking about doing it in the next 6 months (3)
- I am definitely planning to do it in the next 30 days (5)
- I have already done it (6)

ACP4_2 **2. What Matters Most in Life** The following questions are about specific medical treatments that people may or may never want if they were very sick or at the end of their life. For instance, some people know they would want to be on a breathing machine. Other people know they would never want to be on a breathing machine.

**Once again, there are no right or wrong answers and we greatly appreciate your honest opinions.**

**2. How ready are you to talk to your MEDICAL DECISION MAKER about the kind of medical care you would want if you were very sick or near the end of life?**

- I have never thought about it (1)
- I have thought about it, but I am not ready to do it (2)
- I am thinking about doing it in the next 6 months (3)
- I am definitely planning to do it in the next 30 days (5)
- I have already done it (6)

ACP4_3 "A medical provider includes people on your medical care team including doctors, physician assistants, nurse practitioners, nurses, social workers, case managers, and other providers.”

**3. How ready are you to talk to your MEDICAL PROVIDERS about the kind of medical care you would want if you were very sick or near the end of life?**

- I have never thought about it (1)
- I have thought about it, but I am not ready to do it (2)
- I am thinking about doing it in the next 6 months (3)
- I am definitely planning to do it in the next 30 days (5)
- I have already done it (6)

ACP4_4 **4. How ready are you to SIGN OFFICIAL PAPERS putting your wishes about the kind of medical care you would want if you were very sick or near the end of life?**

- I have never thought about it (1)
- I have thought about it, but I am not ready to do it (2)
- I am thinking about doing it in the next 6 months (3)
- I am definitely planning to do it in the next 30 days (5)
- I have already done it (6)
