## Supplementary Appendix Survey Instrument - Caregivers for "Knowledge, Attitudes, and Practices of Advance Care Planning in Patients with Neurosurgical Conditions and Their Caregivers"

#### **Supplementary Appendix Survey Instrument – Caregiver**

### **ACP-NES Study Questionnaire for Caregivers**

### Introduction

This study is titled “Advance Care Planning in Neurology, Neuro-oncology, and Surgery (ACP-NES Study)”. Thank you for taking the time to fill in this online survey. Before doing so, kindly read the consent form to ensure that you understand the study and what is involved.

[Consent form will be in hardcopy form.]

1. Gender

☐ M

☐ F
☐ Others

☐ Prefer Not To Say

2. Age

☐ 21 – 25 years old	☐ 26 – 35 years old	☐ 36 – 45 years old

☐ 46 – 54 years old	☐55 – 64 years old	☐ 46 – 54 years old

☐ ≥ 75 years old

3. Marital status:

☐Single

☐Married

☐Divorced

☐Widowed

4. Education

☐ Primary and Below

☐ Secondary School

☐ Pre-Tertiary (GCE A-Levels, Diploma)

☐ Bachelor’s Degree

☐ Post-Graduate Degree (Master’s or PhD)

5. Race

☐ Chinese

☐ Indian

☐ Malay

☐ Others ___

6. Religion

☐ Buddhism

☐ Christianity

☐ Hinduism

☐ Islam

☐ No Religion

☐ Others ___

7. Socio-economic status
   7.1. Income brackets:

☐ < 30,000 SGD / year

☐ < $30,000 – $49,000 SGD / year

☐ $50,000 – $99,000 SGD / year

☐ > $100, 000 SGD / year

7.2. Housing type:

☐ 1 – 2-room HDB Flat

☐ 3 – 4-room HDB Flat

☐ 5-room and Executive HDB Flat

☐ Condominiums and Other Apartments

☐ Landed Properties

8. What is your relationship to the patient? Ie; “The patient is my…”

☐ Parent
☐ Grandparent
☐ Spouse

☐ Others; please specify:

9. Have you worked in the healthcare field before? (eg; as a nurse, physician, pharmacist etc;)

☐ No
☐ Yes; please specify your most recent position: ______________

10. Have you ever been involved in making end-of-life decisions for someone else in the past?
    ☐ No
    ☐ Yes
11. As a caregiver, you may have to help make medical choices for the patient when the patient is unable to. Currently, how ready do you feel to do this?
    ☐ Very Ready	☐ Somewhat Ready	☐ Neutral
    ☐ Somewhat Not Ready	☐ Not Ready At All
12. Currently, is the patient willing and able to participate in their medical decision making?

☐ Yes the patient is willing and able to.

☐ No, the patient is willing, but unable to participate.

☐ No, the patient is unwilling, but able to participate.

☐ No, the patient is unwilling and unable to participate.

☐ I am not sure.

13. How much do you agree with the following statements?
    13.1. It is easy for the patient and their family to talk about personal and emotional matters.

☐ Completely Disagree

☐ Disagree

☐ Neutral

☐ Agree

☐ Completely Agree

13.2. It is easy for me (a caregiver) and the patient to talk about personal and emotional matters.

☐ Completely Disagree

☐ Disagree

☐ Neutral

☐ Agree

☐ Completely Agree

13.3. I am satisfied with the information I receive from the patient’s doctors

☐ Completely Disagree

☐ Disagree

☐ Neutral

☐ Agree

☐ Completely Agree

13.4. I am well-informed about the possible locations of care for the patient (eg; care at home versus in a nursing home).

☐ Completely Disagree

☐ Disagree

☐ Neutral

☐ Agree

☐ Completely Agree

13.5. The care options available to the patient are limited by my financial situation.

☐ Completely Disagree

☐ Disagree

☐ Neutral

☐ Agree

☐ Completely Agree

14. Has a doctor given the patient a medical prognosis of 12 months or less left to live?

☐ No	☐ Yes	☐ Not sure

15. Has the patient ever expressed their wishes or preferences about their care at the end-of-life?

☐ No	☐ Yes	☐ Not sure

**EQ5D**

**MOBILITY**

- ⊗I have no problems in walking about (1)
- ⊗I have slight problems in walking about (132)
- ⊗I have moderate problems in walking about (133)
- ⊗I have severe problems in walking about (134)
- ⊗I am unable to walk about (135)

**SELF-CARE**

- ⊗I have no problems washing or dressing myself (1)
- ⊗I have slight problems washing or dressing myself (6)
- ⊗I have moderate problems washing or dressing myself (7)
- ⊗I have severe problems washing or dressing myself	(8)
- ⊗I am unable to wash or dress myself (9)

**USUAL ACTIVITIES (e.g. work, study, housework, family or leisure activity):**

- ⊗I have no problems doing my usual activities (6)
- ⊗I have slight problems doing my usual activities (7)
- ⊗I have moderate problems doing my usual activities (8)
- ⊗I have severe problems doing my usual activities (9)
- ⊗I am unable to do my usual activities (10)

**PAIN / DISCOMFORT**

- ⊗I have no pain or discomfort (1)
- ⊗I have slight pain or discomfort (6)
- ⊗I have moderate pain or discomfort (7)
- ⊗I have severe pain or discomfort (8)
- ⊗I have extreme pain or discomfort (9)

**ANXIETY / DEPRESSION**

- ⊗I am not anxious or depressed? (1)
- ⊗I am slightly anxious or depressed? (6)
- ⊗I am moderately anxious or depressed? (7)
- ⊗I am severely anxious or depressed? (8)
- ⊗I am extremely anxious or depressed? (9)

| Page Break |
| --- |

We would like to know how good or bad your health is TODAY.

This scale is numbered from 0 to 100

**100** means the best health you can imagine.

**0** means the worst health you can imagine.

Mark an X on the scale to indicate how your health is TODAY.

|  | **The worst health you can imagine** | **The best health you can imagine** |
| --- | --- | --- |

|  | 0 | 5 | 10 | 15 | 20 | 25 | 30 | 35 | 40 | 45 | 50 | 55 | 60 | 65 | 70 | 75 | 80 | 85 | 90 | 95 | 100 |
| --- | --- | --- | --- | --- | --- | --- | --- | --- | --- | --- | --- | --- | --- | --- | --- | --- | --- | --- | --- | --- | --- |

| () | 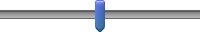 |
| --- | --- |

**Caregiver Reaction Assessment Scale (CRA)**

**Subscale 5: Self-esteem**

| Item | Strongly disagree | Disagree | Neither agree or disagree | Agree | Strongly agree |
| --- | --- | --- | --- | --- | --- |
| 1. Caring for my partner is important to me. | 1 | 2 | 3 | 4 | 5 |
| 2. I enjoy caring for my partner. | 1 | 2 | 3 | 4 | 5 |
| 3. Caring for my partner makes me feel good. | 1 | 2 | 3 | 4 | 5 |
| 4. I feel privileged to care for my partner. | 1 | 2 | 3 | 4 | 5 |
| 5. I resent having to care for my partner (reversed). | 1 | 2 | 3 | 4 | 5 |
| 6. I really want to care for my partner. | 1 | 2 | 3 | 4 | 5 |
| 7. I will never be able to do enough caregiving to repay my partner (reversed). | 1 | 2 | 3 | 4 | 5 |

**Final 17-item ACP Engagement Survey – Surrogate Decision-Maker**

**KNOWLEDGE**

| How well informed are you about….. | Not at all | A little | Somewhat | Fairly | Extremely | Not sure/Ref. |
| --- | --- | --- | --- | --- | --- | --- |
| 1. Who can be a medical decision maker? | 1 | 2 | 3 | 4 | 5 | 8 / 9 |
| 2. What makes someone a good medical decision maker? | 1 | 2 | 3 | 4 | 5 | 8 / 9 |
| 3. The types of decisions that you may have to make for your loved one in the future? | 1 | 2 | 3 | 4 | 5 | 8 / 9 |

**SELF-EFFICACY**

| How confident are you that today you could….. | Not at all | A little | Somewhat | Fairly | Extremely | Not sure/Ref. |
| --- | --- | --- | --- | --- | --- | --- |
| 4. Serve as your loved one’s medical decision maker? | 1 | 2 | 3 | 4 | 5 | 8 / 9 |
| 5. Talk with your loved one about the care he/she would want if he/she were very sick or near the end of life? | 1 | 2 | 3 | 4 | 5 | 8 / 9 |
| 6. Be part of a discussion with your loved one’s DOCTOR about the care your loved one would want if he/she were very sick or near the end of life? | 1 | 2 | 3 | 4 | 5 | 8 / 9 |

**CONTEMPLATION**

| How much have you thought about….. | Never | Once or twice | A few times | Several times | A lot | Not sure/Ref. |
| --- | --- | --- | --- | --- | --- | --- |
| 7. You role as your loved one’s medical decision-maker? | 1 | 2 | 3 | 4 | 5 | 8 / 9 |
| 8. Talking with your loved one about whether or not there are certain health situations that would make your loved one’s life not worth living? | 1 | 2 | 3 | 4 | 5 | 8 / 9 |
| 9. Being part of a discussion with your loved one’s DOCTORS about whether or not there are certain health situations that would make your loved one’s life not worth living? | 1 | 2 | 3 | 4 | 5 | 8 / 9 |
| 10. Talking with your loved one about the care he/she would want if they were very sick or near the end of life? | 1 | 2 | 3 | 4 | 5 | 8 / 9 |
| 11. Being part of a discussion with your loved one’s DOCTORS about the care he/she would want if they were very sick or near the end of life? | 1 | 2 | 3 | 4 | 5 | 8 / 9 |

**READINESS**

12. How ready are you to formally discuss with your loved one your role as their medical decision maker?

□ I have never thought about it

□ I have thought about it, but I am not ready to do it

□ I am thinking about doing it in the next 6 months

□ I am definitely planning to do it in the next 30 days

□ I have already done it

□ Not Sure

□ Refused

OPTIONAL: If they answered, “I have already done it”, then ask “When did you do this?”

□ Less than 6 months ago

□ More than 6 months ago

□ NA

13. How ready are you to talk to your loved one about the kind of medical care he/she would want if they were very sick or near the end of life?

□ I have never thought about it

□ I have thought about it, but I am not ready to do it

□ I am thinking about doing it in the next 6 months

□ I am definitely planning to do it in the next 30 days

□ I have already done it

□ Not Sure

□ Refused

OPTIONAL: If they answered, “I have already done it”, then ask “When did you do this?”

□ Less than 6 months ago

□ More than 6 months ago

□ NA

14. How ready are you to talk to your loved one about whether or not there are certain health situations that would make your loved one’s life not worth living?

□ I have never thought about it

□ I have thought about it, but I am not ready to do it

□ I am thinking about doing it in the next 6 months

□ I am definitely planning to do it in the next 30 days

□ I have already done it

□ Not Sure

□ Refused

OPTIONAL: If they answered, “I have already done it”, then ask “When did you do this?”

□ Less than 6 months ago

□ More than 6 months ago

□ NA

15. How ready are you to be part of a discussion with your loved one’s DOCTOR about whether or not there are certain health situations that would make your loved one’s life not worth living?

□ I have never thought about it

□ I have thought about it, but I am not ready to do it

□ I am thinking about doing it in the next 6 months

□ I am definitely planning to do it in the next 30 days

□ I have already done it

□ Not Sure

□ Refused

OPTIONAL: If they answered, “I have already done it”, then ask “When did you do this?”

□ Less than 6 months ago

□ More than 6 months ago

□ NA

16. How ready are you to be part of a discussion with your loved one’s DOCTOR about the kind of medical care your loved one would want if he/she were very sick or near the end of life?

□ I have never thought about it

□ I have thought about it, but I am not ready to do it

□ I am thinking about doing it in the next 6 months

□ I am definitely planning to do it in the next 30 days

□ I have already done it

□ Not Sure

□ Refused

OPTIONAL: If they answered, “I have already done it”, then ask “When did you do this?”

□ Less than 6 months ago

□ More than 6 months ago

□ NA

17. How ready are you to ask your loved one’s DOCTOR questions to help you make a good medical decision for your loved one if your loved one was unable to speak for themselves?

□ I have never thought about it

□ I have thought about it, but I am not ready to do it

□ I am thinking about doing it in the next 6 months

□ I am definitely planning to do it in the next 30 days

□ I have already done it

□ Not Sure

□ Refused

OPTIONAL: If they answered, “I have already done it”, then ask “When did you do this?”

□ Less than 6 months ago

□ More than 6 months ago

□ NA

**Pearlin's Mastery Scale**

|  | Strongly agree (1) | Agree (2) | Disagree (3) | Strongly disagree (4) |
| --- | --- | --- | --- | --- |
| 1. There is really no way I can solve some of the problems I have |  |  |  |  |
| 2. Sometimes I feel that I’m being pushed around in life |  |  |  |  |
| 3. I have little control over the things that happen to me |  |  |  |  |
| 4. I can do just about anything I really set my mind to |  |  |  |  |
| 5. I often feel helpless in dealing with the problems of life |  |  |  |  |
| 6. What happens to me in the future mostly depends on me |  |  |  |  |
| 7. There is little I can do to change many of the important things in my life |  |  |  |  |

SINGAPORE CAREGIVER QUALITY OF LIFE SCALE 10-ITEM VERSION 1.0 (SCQOLS-10)

Below is a list of statements that other people providing care to sick family members consider important. **For each statement, please circle one answer that best describes your experience in providing care to your sick family member in the past seven days**. There is no right or wrong answer. It is just about your own experience.

PW **Physical Well-being**

|  | Not at all (1) | A little (2) | Somewhat (3) | Quite a lot (4) | Very much (5) |
| --- | --- | --- | --- | --- | --- |
| I have had more aches and pains due to caregiving activities (DCS_1) |  |  |  |  |  |
| My appetite is poor (DCS_2) |  |  |  |  |  |

MW **Mental Well-being**

|  | Not at all (1) | A little (2) | Somewhat (3) | Quite a lot (4) | Very much (5) |
| --- | --- | --- | --- | --- | --- |
| I am fearful of losing my sick family member (DCS_1) |  |  |  |  |  |
| I feel frustrated (DCS_2) |  |  |  |  |  |

EM **Experience & Meaning**

|  | Not at all (1) | A little (2) | Somewhat (3) | Quite a lot (4) | Very much (5) |
| --- | --- | --- | --- | --- | --- |
| I am thankful for the good things that have happened to me even in difficult times (DCS_1) |  |  |  |  |  |
| Caring for my sick family member has drawn the family closer together (DCS_2) |  |  |  |  |  |

DL **Impact on Daily Life**

|  | Not at all (1) | A little (2) | Somewhat (3) | Quite a lot (4) | Very much (5) |
| --- | --- | --- | --- | --- | --- |
| I am bothered that I am not able to leave home or hospital (DCS_1) |  |  |  |  |  |
| I am not able to do what I want to do (DCS_2) |  |  |  |  |  |

FW **Financial Well-being**

|  | Not at all (1) | A little (2) | Somewhat (3) | Quite a lot (4) | Very much (5) |
| --- | --- | --- | --- | --- | --- |
| I am troubled that my sick family member's condition is depleting my savings (DCS_1) |  |  |  |  |  |
| I feel uncertain about my future financial situation (DCS_2) |  |  |  |  |  |
